# Comparing Traditional and Newer Definitions of Obesity in Relation to Incident Cardiovascular Disease and Obesity-Related Cancer Risk: A Prospective Cohort Study in ARIC

**DOI:** 10.1101/2025.11.17.25340450

**Authors:** Omar M Makram, Viraj Shah, Tarek Nahle, Xiaoling Wang, Ryan A Harris, Steven S. Coughlin, Neal L Weintraub, Corinne E. Joshu, Elizabeth A. Platz, Avirup Guha

**Author notes:** Correspondence to: Avirup Guha; Division of Cardiology, Department of Medicine, Medical College of Georgia at Augusta University, Augusta, GA 30912, USA. Final manuscript authors (Elizabeth A. Platz, Avirup Guha). ARIC Senior author.

## Abstract

**Background:** Obesity is a risk factor for both cardiovascular disease (CVD) and cancer. The traditional body mass index (BMI)-based obesity definition (BMI ≥30 kg/m²) has limitations across adulthood and ancestry. The Global Commission on Clinical Obesity in 2025 proposed a new definition incorporating central adiposity measures. The magnitude of increased disease risk among the newly classified obese persons remains undetermined. We compared the associations between obesity definitions and incident CVD and obesity-related cancer in a community-based cohort.

**Methods:** We analyzed 14,834 Atherosclerosis Risk in Communities (ARIC) study participants followed for a median of 25 years. Participants were classified as obese using the traditional definition (BMI ≥30 kg/m²) and the new definition (BMI ≥25 kg/m² plus ≥1 central adiposity measure, or two central adiposity measures as follows: waist circumference ≥102 cm Male/ ≥88 cm Female; waist-to-hip ratio >0.90 Male/ >0.85 Female; waist-to-height ratio >0.5). We identified participants classified as obese only by the new definition. Cox models adjusted for demographics, socioeconomic, and clinical risk factors estimated the adjusted hazard ratios (aHR). A subgroup analyses by age, sex, and race, and time-dependent sensitivity analyses were conducted.

**Results:** 54% and 27% were classified as obese, respectively, according to the new-only (median BMI 26 kg/m^2^) versus the traditional (median BMI 33 kg/m^2^) definition; 19% were non-obese by both. 7% of the “obese by new-only definition” group (i.e. BMI 25-<30 kg/m^2^ and central adiposity) reverted to non-obese status. Compared to non-obese, both new-only (aHR 1.25, 95% CI 1.15-1.35) and traditionally-defined (aHR 1.65, 95% CI 1.51-1.81) obesity were associated with higher CVD risk. The same pattern was noted in coronary heart disease and heart failure. For obesity-related cancers, the traditional definition (aHR 1.36, 95% CI 1.16-1.59), but not the new-only definition (aHR 1.09, 95% CI 0.95-1.26), conferred significantly higher risk. The traditional definition was also associated with higher risk of total and colorectal cancers. Time-dependent analysis and subgrouping by sex, race, and age at time of diagnosis yielded similar results.

**Conclusion:** Both obesity definitions identified increased CVD risk, with significant trends across groups. Only traditionally-defined obesity consistently showed increased risk for total and obesity-related cancers. The newer, more sensitive, definition identified a large cohort with intermediate CVD risk, not previously captured by BMI alone, highlighting a distinct risk profile, suggesting potential for targeted interventions. These findings warrant careful consideration of potentially inflated risk in patients defined as obese by the new criteria.

**Funding:** NHLBI, NCI, NPCR, AHA

## Introduction

Obesity is a major global health challenge and a significant risk for several conditions, primarily cardiovascular diseases (CVD) and various types of cancer.^1,2^ Traditionally, obesity screening and definition have relied heavily on the body mass index (BMI), with a threshold of ≥30 kg/m² frequently used for classification. While convenient and widely accepted for population surveillance and initial screening, its limitations as an individual-level measure of health risk are increasingly recognized.^3^ BMI does not directly measure body adiposity, differentiate between lean mass and fat mass, or account for the body fat distribution, particularly central adiposity, which reflects visceral adiposity and is considered more detrimental.^4^ Central adiposity, often quantified through measures like waist circumference (WC), waist-to-hip ratio (WHR), and waist-to-height ratio (WHTR), reflects visceral fat accumulation and is increasingly considered a stronger predictor of cardiometabolic risk and mortality than BMI alone,^5–7^ suggesting that BMI may misclassify risk by overestimating it in those with high lean mass, such as athletes, or by underestimating it in those with normal BMI but high central adiposity.^3^ Some studies of abdominal obesity have suggested that it may play a role in cancer development or progression,^8–10^ but results to date have been somewhat inconsistent and additional research is needed. Central obesity promotes low grade chronic inflammation that increases tumor-promoting oxidative stress.^11^ More importantly, BMI does not necessarily directly correlate with the extent of adiposity and lean muscle mass across adulthood and various ancestry groups.^12^ Recognizing these limitations, newer frameworks, such as those proposed by the Lancet Commission report, advocate for combining BMI with central adiposity measures to more effectively identify high-risk adiposity patterns that BMI alone may miss, and to classify obesity based on organ dysfunction into preclinical and clinical obesity.^3^

Despite the recognized limitations of BMI usage alone and the proposed importance of central adiposity, to our knowledge, no previous large-scale prospective studies have directly compared the long-term association between obesity according to the new versus the old definition and CVD and cancer risk. Moreover, the potential impact of pre-existing conditions, such as baseline CVD or history of cancer, on these associations is not fully understood. Addressing these gaps and nuances is critical for refining risk stratification and tailoring prevention strategies.

In this study, we aimed to address these gaps by comparing the traditional BMI-based definition of obesity with the newer definition that incorporates central adiposity measures along with BMI to predict the incidence of CVD and cancer in a large, community-based prospective cohort of female and male, Black and White participants.

## Methods

### Study Design and Population

We utilized the Strengthening the Reporting of Observational Studies in Epidemiology (STROBE) cohort checklist to report the outcomes **(Supplemental material, STROBE Statement)**.^13^ The Atherosclerosis Risk in Communities (ARIC) study was approved by the institutional review boards (IRBs) at all participating institutions and all participants provided written informed consent at each study visit. Data is available upon request from ARIC.

In this analysis, we used data from the ARIC study, which included individuals aged 45 to 64 years at the beginning of follow-up. We included participants who started follow-up at visit 1 (1987-1989) from the four US communities used in the ARIC study (Forsyth County, North Carolina; Jackson, Mississippi; suburban Minneapolis, Minnesota; and Washington County, Maryland). Participants were followed up every three years for the first four visits as follows: visit 2 (1990-1992), visit 3 (1993-1995), visit 4 (1996-1998), then there was a gap until visit 5 (2011-2013). Several other visits continued as follows: visit 6 (2016-2017), visit 7 (2018-2019), visit 8 (2020), visit 9 (2021-2022), visit 10 (2023), and visit 11 (2024-2025). This study used data from visits 1 through visit 5 (for cancer outcomes) and visit 10 (for cardiovascular outcomes). The current analysis included 14,834 participants who attended visit 1, had available obesity measurements (BMI, waist circumference, waist-to-hip ratio, and waist-to-height ratio), and provided informed consent. Visit 1 was designated as the start of follow-up, and participants were monitored from visit 1 through December 31, 2015, for cancer development and through December 31, 2023, for cardiovascular outcomes, or until the occurrence of the outcome, loss to follow-up, or administrative censoring, whichever came first. As ARIC study is still ongoing, administrative censoring was defined as December 31, 2015, for cancer development and December 31, 2023, for cardiovascular outcomes. For individuals who did not develop any outcome, they were censored at the time of death if died, or at the administrative censoring date if still alive. We excluded individuals with a history of cancer (n=974) at or before the beginning of follow-up when examining the incidence of cancer. Similarly, we excluded individuals with a history of CVD (n=2,481) at or before the beginning of follow-up when examining the incidence of CVD **(Figure 1)**.

**Figure 1.**
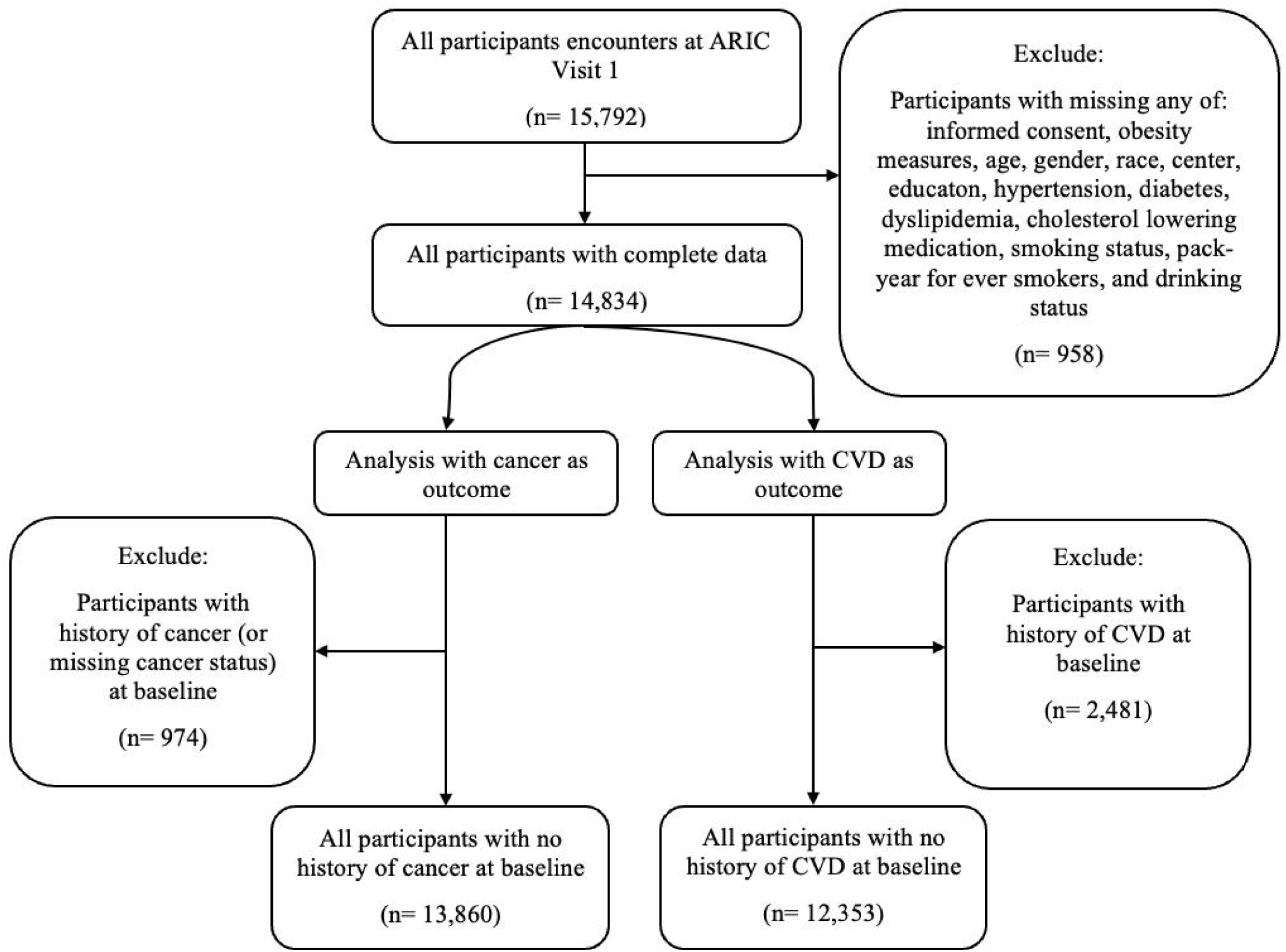
Flow chart for the included participants in the analysis for both CVD and cancer outcomes.

### Exposure Assessment

The primary exposure was the obesity definition used as a categorical variable estimated using the baseline measurements of BMI, WC, WHR, and WHTR, and individuals were categorized as follows: 1) those identified by the new definition of obesity only; 2) those identified as being obese by both definitions or by the old definition only; 3) individuals identified as non-obese by both definitions (BMI < 30 kg/m² and adiposity thresholds below those described below). The first group was defined as obese based on the new definition as follows: (1) BMI ≥ 25 kg/m² AND at least one measurement of body size above the threshold (WC ≥ [threshold] cm OR WHR > [threshold] OR WHTR > [threshold]); (2) having at least two of measurements of body size above threshold, regardless of BMI; (3) BMI > 40 kg/m². Standing height, waist girth, and hip girth was measured to the nearest centimeter. Weight was measured to the nearest pound (lb). All numbers were rounded down to the nearest whole number. For BMI calculation, the following formula was used: [weight (lbs) / 2.20] / [height (cm) / 100]^2^ Thresholds were based on standard gender-specific cutoffs as described in the recent Lancet Commission report, e.g., WC ≥ 102 cm for males/ ≥ 88 cm for females; WHR > 0.90 for males/> 0.85 for females; WHTR > 0.50.^3^ The older definition used a BMI of 30 kg/m² or above for obesity.

### Outcome: Incidence of CVD and Cancer

The first primary outcome, incident CVD, was defined as a composite of the first event of (1) coronary heart disease (CHD), defined as a definite or probable myocardial infarction, definite fatal coronary heart disease, or coronary revascularization procedure;^14^ (2) heart failure (HF), defined as hospitalization or death from heart failure with the International Classification of Diseases Code, Ninth Revision (ICD-9) 428 or Tenth Revision (ICD-10) I50;^15^ (3) definite or probable stroke or transient ischemic attack (TIA), defined as having sudden or rapid onset of neurological symptoms lasting for 24 hours (or less than 24 hours for TIA) or leading to death, using previously established methods.^16,17^

The second primary outcome was incident obesity-related cancer (post-menopausal breast, colorectal, liver, pancreas, endometrial, ovarian, kidney, or thyroid).^2,18^ The secondary outcomes included specific incident specific cardiovascular outcomes (CHD, HF, and stroke/TIA), primary total cancer, and site-specific cancers. Site-specific cancers included prostate cancer in males, post-menopausal breast cancer in females, lung cancer, colorectal cancer, kidney cancer, hematopoietic/lymphatic cancers, and any cancer except prostate. Any cancer, except prostate cancer, was considered separately because prostate cancer is not known to be directly related to obesity, so we aimed at investigating the full impact of obesity on cancer without the attenuated effect from prostate cancer.^2^ Cancer events were identified through the linkage to the state cancer registries in North Carolina, Mississippi, Minnesota, and Maryland from 1987 to 2015. Medical records, hospital discharge summaries, and codes, as well as death certificates, further supplemented this.^19^

### Covariate Assessment

We used demographic, socioeconomic variables, and CVD and/or cancer risk and protective factors variables collected at baseline. Demographic information included age, sex (male, female), race (White, Black), and participating center. For this analysis, another variable (race*center) was created as follows NC-White, NC-Black, MS-Black, MN-White, MD-White. Race and center were tightly linked; for instance, Jackson, MS center was restricted to Black individuals only during recruitment. Socioeconomic covariates included education level, health insurance, and frequency of routine physical examination. CVD and/or cancer risk and protective factors variables included hypertension, defined as systolic blood pressure ≥140 mmHg, diastolic blood pressure ≥ 90 mmHg, the use of antihypertensive medications, or reporting taking antihypertensive medications within the last 2 weeks; diabetes, defined as fasting glucose ≥ 126 mg/dL, non-fasting glucose ≥ 200 mg/dL, self-reported physician diagnosis, or use of diabetes medications in the last 2 weeks; dyslipidemia, assessed through standard lipid measurements (low density lipoprotein [LDL] ≥ 160 mg/dL or 4.1 mmol/L or triglycerides > 150 mg/dL or 1.7 mmol/L)^20^ or reporting the use of statin or cholesterol-lowering medications within the last two weeks; smoking status (never, former, current) and pack-years of cigarette smoking in those who ever smoked; alcohol consumption (current, former, never); hormone replacement therapies in females (current, former, never); chronic kidney disease (CKD), defined using the estimated glomerular filtration rate (eGFR) using the creatinine equation.

### Statistical Analysis

An unadjusted baseline characteristics comparison was conducted for all included categorical and continuous variables using frequencies and proportions, as well as medians and interquartile intervals (IQI, i.e., 25-75 percentiles). Formal statistical tests were conducted using chi-square tests for categorical variables and Kruskal-Wallis tests for continuous variables. As a sensitivity analysis, we also include a supplementary table for baseline characteristics using a BMI of ≥30 kg/m^2^ in the new definition, the same cutoff used in the BMI-based definition.

After confirming the proportional hazards assumption using the Schoenfeld residuals, we used Cox proportional hazards models to estimate cause-specific hazard ratios (HR) and the 95% confidence intervals (95% CI) for the association between the obesity definition groups and the incidence of CVD and cancer. Each outcome analysis was conducted independently; participants could contribute to both analyses if they were free of both conditions at baseline. In these models, participants who died without experiencing the outcome of interest were censored at the time of death. This approach estimated the hazard of developing the outcome among participants who remain alive and event-free, which was deemed appropriate for understanding etiologic associations between obesity definitions and disease incidence. We employed sequential modeling using a priori set of covariates as follows: Minimally adjusted model (Model 0): adjusted for age, sex, and race*center; Model 1: Model 0 + education; Model 2: Model 1 + CVD and/or cancer risk and protective factors (hormonal therapy in females [Male, Female-no HRT, Female HRT], hypertension, diabetes, dyslipidemia, statin or cholesterol-lowering medication use, history of smoking, smoking pack-years for ever smokers, and history of drinking). We noticed that the minimally adjusted model (model 0) and model 1 were very similar for all outcomes, so we decided to report models 1 and 2 only for clarity. P-trend analysis for our primary categorical exposure was calculated by assigning ordered numerical values to its groups and including this single variable as a continuous, linear term in the Cox proportional hazards models and reporting its p-value.

Our primary analyses used obesity status and covariates measured at baseline. As a sensitivity analysis, we used the obesity definition variable as a time-dependent (time-updated) variable to incorporate the obesity estimation calculated at each visit and allow for obesity status to be updated at each visit (e.g., a participant identified as non-obese at visit 1 might become obese at visit 3 according to the old definition). A time-updated Cox regression model, using Lexis expansion, was conducted to split the follow-up time into visit-defined intervals, updating obesity status at each visit.^21^

Subgrouping by sex, race, and age was further conducted for all the outcomes to address the BMI variability across adulthood and ancestry. For age subgrouping, we used the median age at diagnosis observed in those who experienced the outcome as the cutoff. This was conducted because advancing age is commonly associated with changes in body composition including loss of height, decreased lean muscle mass and increased fat percentage, which may modify the relationship between obesity definition and the outcome of CVD or cancer.^22^

Finally, a complete case analysis approach was used for the main analysis. However, to ensure robustness, a multiple imputation by chained equations (MICE) approach was employed, addressing the minimal missingness (less than 2%) observed in the covariates. Twenty datasets were generated (M=20), and the final model estimates were pooled using the Rubin’s rules.^23^ Only the height at the second visit was not recorded; thus, we used the height from the first visit instead. All other visits had heights and other anthropometric measures recorded. All statistical analysis was conducted using STATA/IC 16.1 (StataCorp, College Station, TX, USA), and two-sided p-values <0.05 were deemed statistically significant.

## Results

### Baseline characteristics

We included 14,834 eligible participants at visit 1 **(Figure 1)**, with a median age of 54 years (IQI 49-59 years). Females and Blacks comprised about 54% and 26% of the analytic cohort, respectively. We identified 19% (n=2,834) of the participants being non-obese by both definitions, 54% (n=7,953) of the participants were newly identified as obese by the new-only definition, and about 27% (n=4,047) defined as obese by either the older definition only (n=3) or both definitions (n=4,044; **Table 1)**. In the sensitivity analysis, when using a BMI cutoff of ≥30 kg/m^2^ in the new definition instead of 25 kg/m^2^, a similar pattern was noted, with about 51% of the total population identified as obese by the new definition. A total of 448 (3%) participants had a BMI between 25 and 30 kg/m^2^ and had only one of the abdominal obesity measurements above the identified thresholds; thus, they were defined as non-obese as the BMI threshold was adjusted to 30 kg/m^2^ **(Table S1)**.

**Table 1.** Baseline clinical characteristics of participants by obesity definitions. Bold text indicates significant results.

| **Characteristics** | **Total** | **Non-Obese by both**<br>**definitions** | **Obese by new**<br>**definition only** | **Obese by both or**<br>**old definitions** |
| --- | --- | --- | --- | --- |
| **Sample size** | 14,834 | 2,834 (19%) | 7,953 (54%) | 4,047 (27%) |
| **ARIC Center** |  |  |  |  |
| Forsyth, NC | 3,817 (25.7%) | 29.2% | 28.7% | 17.5% |
| Jackson, MS | 3,349 (22.6%) | 21.0% | 17.2% | 34.2% |
| Minneapolis, MN | 3,871 (26.1%) | 32.0% | 26.3% | 21.6% |
| Washington, MD | 3,797 (25.6%) | 17.7% | 27.9% | 26.7% |
| **Age** | 54.0 (49.0-59.0) | 52.0 (48.0-57.0) | 55.0 (50.0-60.0) | 54.0 (49.0-59.0) |
| **Gender** |  |  |  |  |
| Male | 6,776 (45.7%) | 35.7% | 52.4% | 39.4% |
| Female | 8,058 (54.3%) | 64.3% | 47.6% | 60.6% |
| **Race** |  |  |  |  |
| White | 11,029 (74.3%) | 76.3% | 80.0% | 61.9% |
| Black | 3,805 (25.7%) | 23.7% | 20.0% | 38.1% |
| **Current/former HRT in females** | 2,721 (33.8%) | 38.1% | 36.4% | 26.5% |
| **Social determinants of health** |  |  |  |  |
| **Education** |  |  |  |  |
| No or below 12 years of education | 3,466 (23.4%) | 15.5% | 22.6% | 30.3% |
| High school education | 6,069 (40.9%) | 41.2% | 41.4% | 39.7% |
| At least some college | 5,299 (35.7%) | 43.3% | 36.0% | 29.9% |
| **Health insurance** |  |  |  |  |
| Yes | 13,397 (90.4%) | 91.8% | 91.9% | 86.5% |
| No | 1,423 (9.6%) | 8.2% | 8.1% | 13.5% |
| **Frequency of Physical Examination (PE)** |  |  |  |  |
| At least once a year | 6,396 (43.2%) | 46.0% | 41.5% | 44.5% |
| At least once every five years | 3,568 (24.1%) | 23.9% | 25.5% | 21.4% |
| Less than once every five years | 681 (4.6%) | 3.8% | 4.8% | 4.7% |
| Do not have routine PE | 4,170 (28.1%) | 26.4% | 28.1% | 29.4% |
| **Anthropometrics** |  |  |  |  |
| **Waist circumference, cm** | 96.0 (88.0-105.0) | 80.0 (75.0-84.0) | 95.0 (90.0-100.0) | 111.0 (105.0-118.0) |
| **Waist hip ratio** | 0.9 (0.9-1.0) | 0.8 (0.8-0.9) | 0.9 (0.9-1.0) | 1.0 (0.9-1.0) |
| **Waist height ratio** | 0.6 (0.5-0.6) | 0.5 (0.5-0.5) | 0.6 (0.5-0.6) | 0.7 (0.6-0.7) |
| **Height, cm** | 168.0 (161.0-176.0) | 167.0 (162.0-174.0) | 169.0 (162.0-176.0) | 167.0 (161.0-174.0) |
| **Body Mass Index (BMI), kg/m2** | 26.9 (24.0-30.4) | 22.2 (20.7-23.6) | 26.4 (24.7-28.0) | 33.1 (31.3-36.3) |
| **BMI category by old definition** |  |  |  |  |
| Under Weight | 136 (0.9%) | 4.6% | 0.1% | 0.0% |
| Healthy Weight | 4,797 (32.3%) | 89.7% | 28.4% | 0.0% |
| Overweight | 5,854 (39.5%) | 5.7% | 71.5% | 0.0% |
| Obesity Class I | 2,716 (18.3%) | 0.0% | 0.0% | 67.1% |
| Obesity Class II | 903 (6.1%) | 0.0% | 0.0% | 22.3% |
| Obesity Class III | 428 (2.9%) | 0.0% | 0.0% | 10.6% |
| **CV risk factors** |  |  |  |  |
| **Drinking Status** |  |  |  |  |
| Never Drinker | 3,677 (24.8%) | 21.0% | 22.0% | 33.0% |
| Current drinker | 8,336 (56.2%) | 63.9% | 59.2% | 44.8% |
| Former Drinker | 2,821 (19.0%) | 15.1% | 18.8% | 22.1% |
| **Hypertension** | 5,828 (39.3%) | 23.6% | 36.4% | 55.9% |
| **Diabetes** | 1,129 (7.6%) | 2.0% | 6.3% | 14.2% |
| **Dyslipidemia** | 13,933 (93.9%) | 91.2% | 94.5% | 94.7% |
| **Used statin in last 2 weeks** | 438 (3.0%) | 2.0% | 3.3% | 3.0% |

| Smoking Status |  |  |  |  |
| --- | --- | --- | --- | --- |
| Never Smoker | 6,208 (41.8%) | 40.6% | 39.2% | 48.0% |
| Current Smoker | 3,866 (26.1%) | 33.4% | 26.9% | 19.2% |
| Former Smoker | 4,760 (32.1%) | 25.9% | 33.9% | 32.8% |
| Smoking Pack-years in ever smokers<sup>#</sup> | 24.8 (11-39) | 23 (9.8-37) | 25.5 (12-40) | 23.3 (9.8-38) |

CKD: chronic kidney disease; CV: cardiovascular; CVD: cardiovascular disease; HRT: hormone replacement therapy (male, female not on therapy, female on therapy); NC: North Carolina; MD: Maryland; MN: Minnesota; MS: Mississippi. Categorical variables presented as n (%); continuous variables presented as median (25-75%tile). <sup>#</sup>represents those who are current and former smokers.

Those defined as obese by the new definition were 52% males and 80% White, compared to 39% males and 62% White for those defined as obese by the old definition. We also noted higher levels of high school education or above among participants defined as obese by the new (77%) compared to the old (70%) definition.

There was a higher prevalence of being ever drinkers (78% vs. 67%) and ever smokers (61% vs. 52%), but a lower prevalence of hypertension (36% vs. 56%) and diabetes (6% vs 14%), in those defined as obese by the new compared to the old definition **(Table 1)**. Over the years of follow-up, the proportion of individuals identified as non-obese by the old criteria tended to decrease, while the proportion of those defined as obese tended to increase **(Figure S1)**. In contrast, most individuals classified as obese by the new criteria remained categorized in that group (77% from visit 1 to visit 2), while 8% transitioned to the BMI-based obesity category, and approximately 7% crossed over to the non-obese category. However, of those with a BMI ≥30 kg/m^2^, ∼ 80% stayed in the same category, 9% transitioned to obesity by the new definition, and only 0.2% crossed over to the non-obese category **(Figure 2)**. We also noted that those who were defined as obese by the new definition in visit 1 and transitioned to non-obese in visit 2 had lower median BMI (visit 1: 26.4 kg/m^2^; visit 2: 24.1 kg/m^2^) and waist circumference (visit 1: 95 cm; visit 2: 89 cm) between visits. This same pattern of lower BMI and waist circumference was observed in all the transitions between visits.

**Figure 2.**
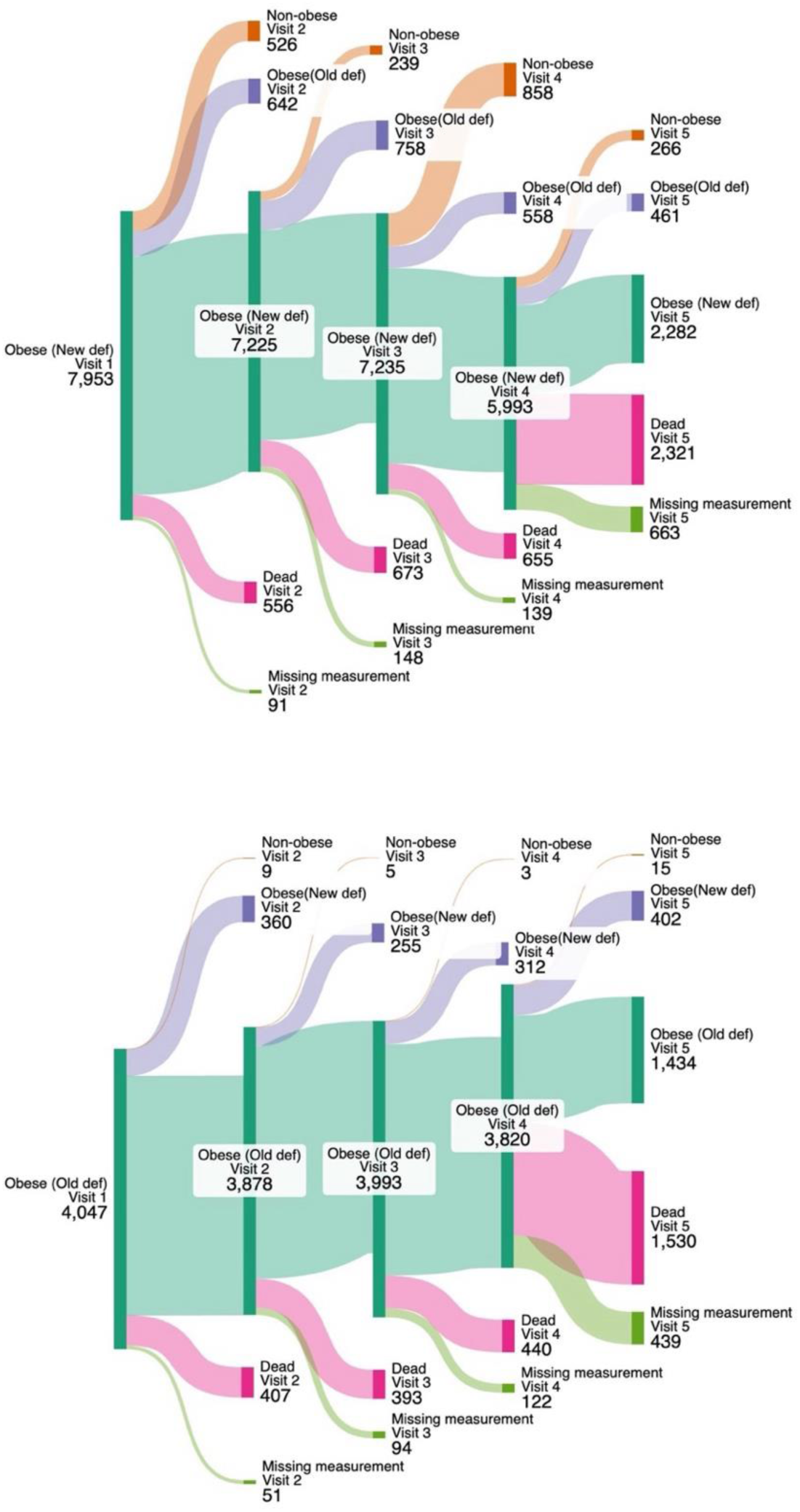
Flow diagram of those defined as obese using the newer and older definitions at baseline.* *The numbers, e.g. for obese (new definition) plus other numbers at visit 2 may not add up to the same number of obese (new definition) at visit 1 because of the transition from other categories such as non-obese or obese (old/both definition) may occur between visits.

### Obesity and CVD

After excluding individuals with a history of CVD at baseline, we followed the participants for a median of 24 years (IQI 14-33 years), during which 5,185 individuals developed CVD. Over the follow-up period, non-obese individuals developed CVD at the rate of 12 CVD per 1,000 cases, followed by those defined as obese by the new-only definition (rate 19 CVD per 1,000 person-years) and those defined as obese by the old/both definition (rate 24 CVD cases per 1,000 person-years). In all sequentially-adjusted models, compared with those considered non-obese by both definitions, those categorized as obese by the new-only definition (Model 2: adjusted hazard ratio [aHR] 1.25, 95% CI 1.15-1.35), or by the old/both definitions (aHR 1.65, 95% CI 1.51-1.81), had a statistically significant higher risk of total CVD and with a significant trend (p<0.001; **Figure 3)**. Yet, the association was weaker among those categorized obese by the new-only definition. A similar pattern was noted with regard to CHD and was even more prominent in HF, where those identified as obese by the new-only definition had a significantly higher risk of heart failure than non-obese individuals (Model 2: aHR 1.28, 95% CI 1.15-1.43). Those identified as obese using the new-only definition did not show significant association with stroke/TIA (Model 2: aHR 0.97, 95% CI 0.83-1.13). A similar finding was noticed in those identified as obese using the old/both definitions, where they showed no significant association with stroke/TIA (Model 2: aHR 1.12, 95% CI 0.95-1.33; **Table 2).** Using the time-updated obesity, the findings were similar for all cardiovascular outcomes, except where those defined as obese using the old/both definitions had significant association with stroke/TIA (aHR 1.24, 95% CI 1.01-1.53**; Table S2)**.

**Figure 3.**
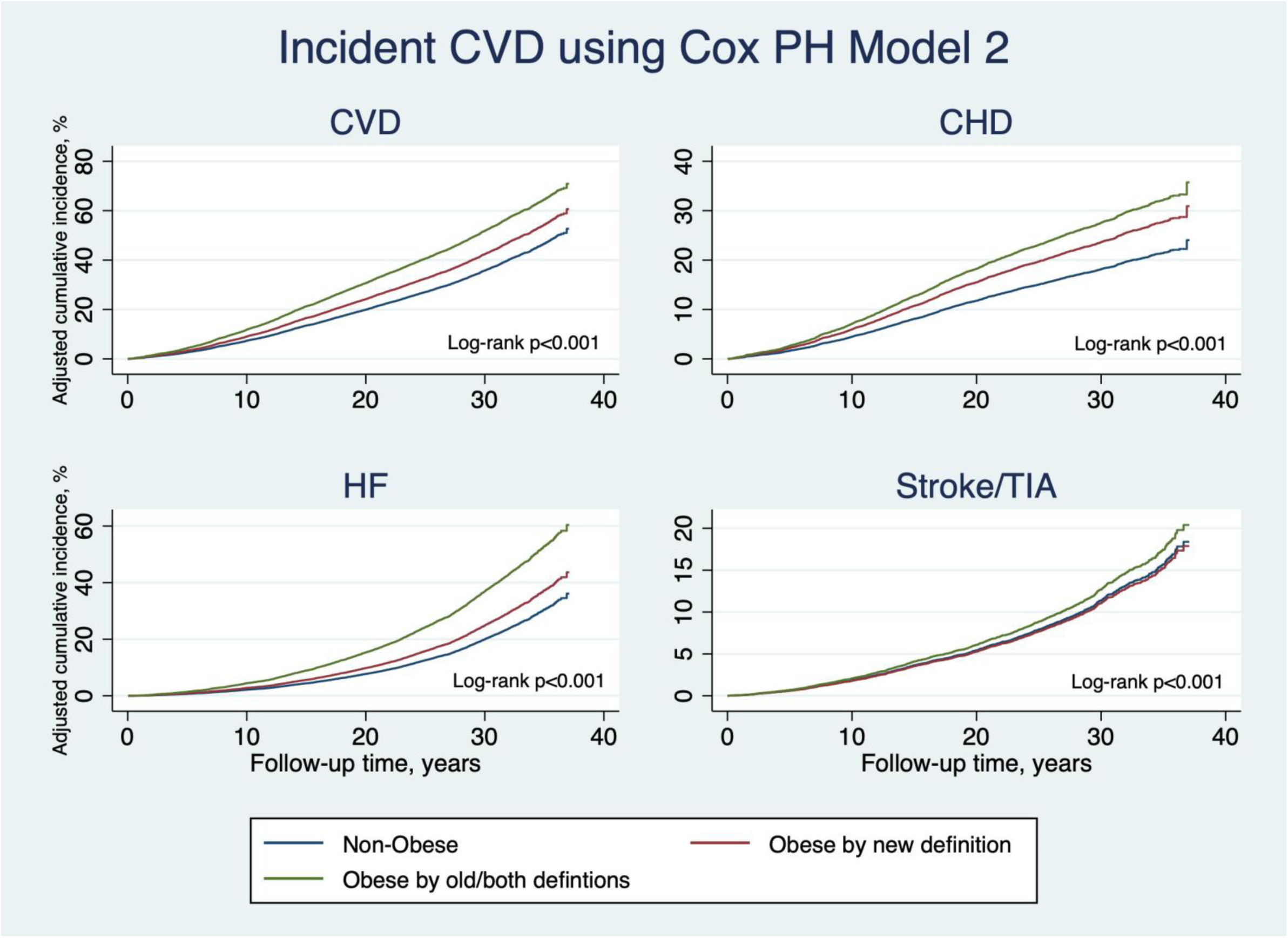
CVD events over time stratified by obesity definition.

**Table 2.**
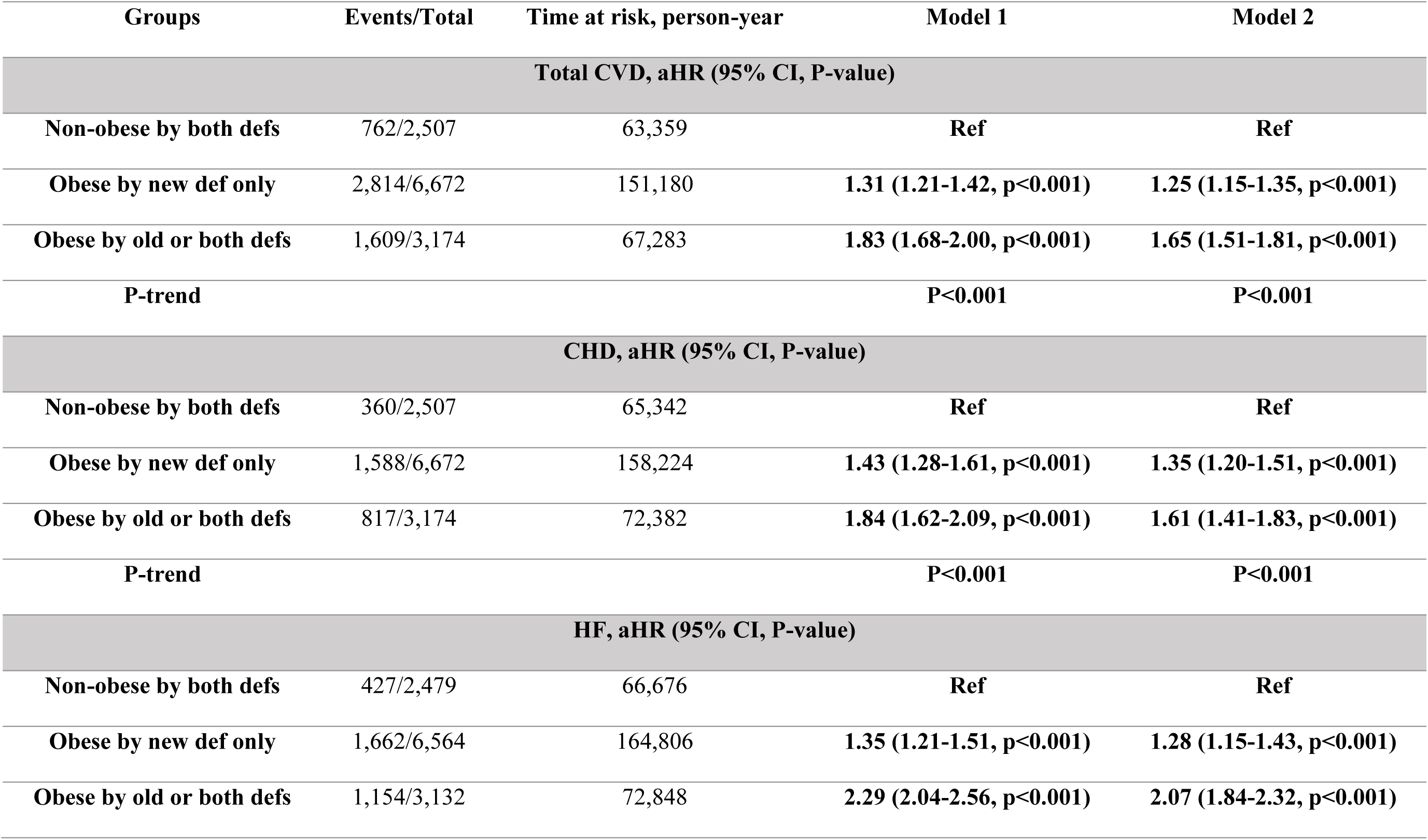
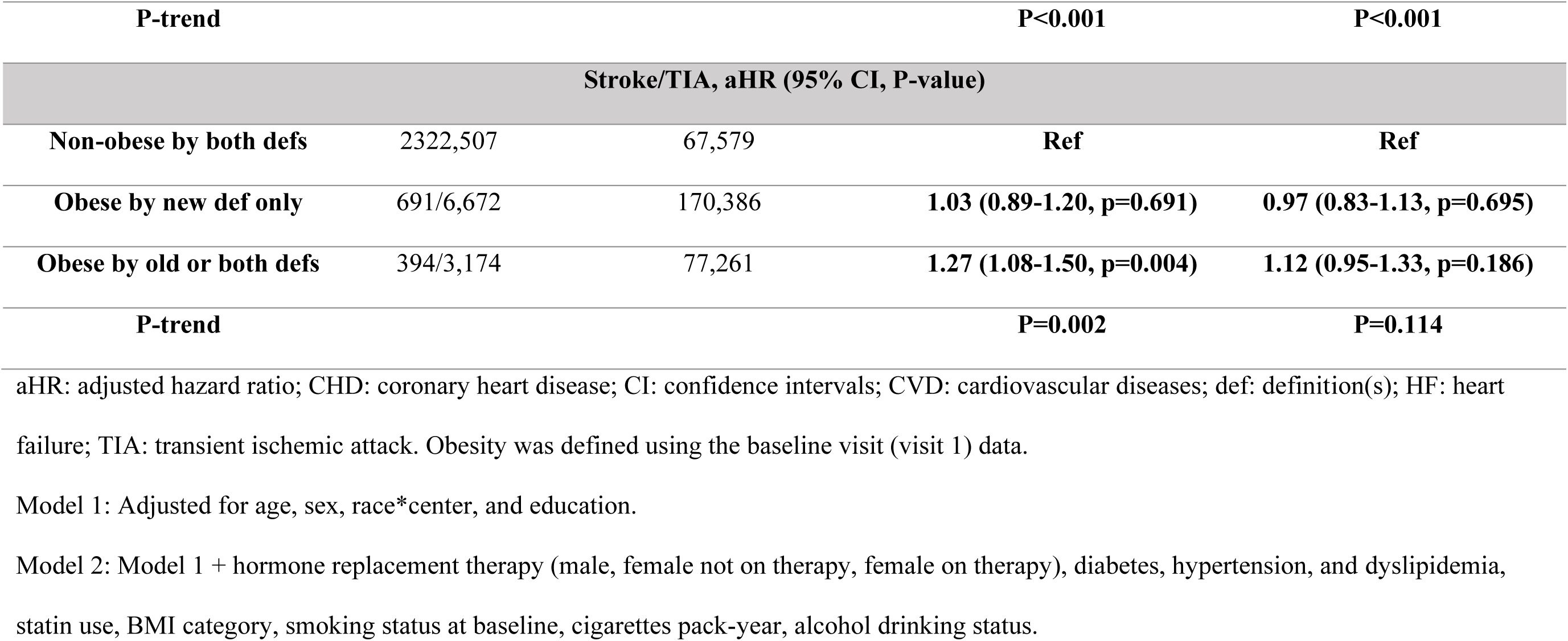
Association between obesity definitions and the incidence of CVD, 12,353 men and women in ARIC, 1987-2015. Bold text indicates significant results.

In the subgroup analysis, both male (n=5,680) and female (n=6,673) subgroups showed similar patterns with a statistically significant association between the new-only obesity definition and total CVD (male: aHR 1.28, 95% CI 1.13-1.44; female: aHR 1.22, 95% CI 1.09-1.36), and a stronger association between the old/both definitions and total CVD (male: aHR 1.58, 95% CI 1.38-1.81; female: aHR 1.72, 95% CI 1.52-1.95). In the race subgroups a very similar pattern was observed, where Whites (n=9,228) showed higher total CVD incidence with both obesity definitions, yet the new-only definition had an attenuated effect (new-only definition: aHR 1.23, 95% CI 1.11-1.35; old/both definitions: aHR 1.64, 95% CI 1.47-1.83; **Table S3**). Upon further subgrouping by race/gender, the same pattern emerged, with White Males and White females exhibiting the same pattern of associations **(Table S4)**. For Blacks (n=3,125), while both definitions showed the same pattern, upon further sub-stratifying into Black males and Black females, both groups demonstrated a statistically significant increase using the old/both definitions but not with the new-only definition **(Table S4)**. It was, however, notable that with HF, both definitions were able to recognize the higher risk of HF across sex and race subgroups **(Tables S5-S6)**.

Lastly, both subgroups of individuals who had events or censored below or above the age of 70 had similar patterns, where those defined as obese using the new-only definition and the old/both definitions had higher risk for CVD compared to non-obese **(Table S3)**. A very similar pattern of associations was noted with other cardiovascular outcomes, where those identified as obese by the old/both definitions had a significant association with the outcome and those identified as obese using the new-only definition had either a significant, yet weaker, association or a non-significant association **(Tables S7-S8)**.

### Obesity and Cancer

After excluding individuals with a history of cancer at baseline, we followed the participants for a median of 25 years (IQI 15-27 years), during which 4,390 individuals were diagnosed with a first primary cancer. Compared with non-obese by both definitions, those categorized as obese by the old/both definitions had a higher risk of total cancer (Model 2: aHR 1.13, 95% CI 1.02-1.24), whereas those categorized as obese by the new-only definition did not have a higher risk of cancer (aHR 1.02, 95% CI 0.93-1.10). The association for obesity categorized by the old/both definitions was stronger for obesity-related cancer (aHR 1.36, 95% CI 1.16-1.59) than for total cancer and was substantially weaker and not statistically significant for obesity categorized by the new-only definition (aHR 1.09, 95% CI 0.95-1.26; **Figure 4**, **Table 3)**. A similar pattern was observed in colorectal cancer (old/both definitions: aHR 1.41, 95% CI 1.03-1.92; new-only definition: aHR 0.99, 95% CI 0.74-1.31). Only for endometrial and kidney cancers did both definitions show a higher risk (old/both definitions: aHR 2.41, 95% CI 1.56-3.72; new-only definition: aHR 1.84, 95% CI 1.23-2.76). On the other hand, both definitions of obesity showed a lower risk of lung cancer (old/both definitions: aHR 0.77, 95% CI 0.60-0.98; new-only definition: aHR 0.80, 95% CI 0.66-0.98). No significant association between obesity, whether by old/both definitions or the new-only definition, and the incidence of post-menopausal breast cancer, prostate cancer, or hematopoietic and lymphatic cancer was observed **(Table 3)**.

**Figure 4.**
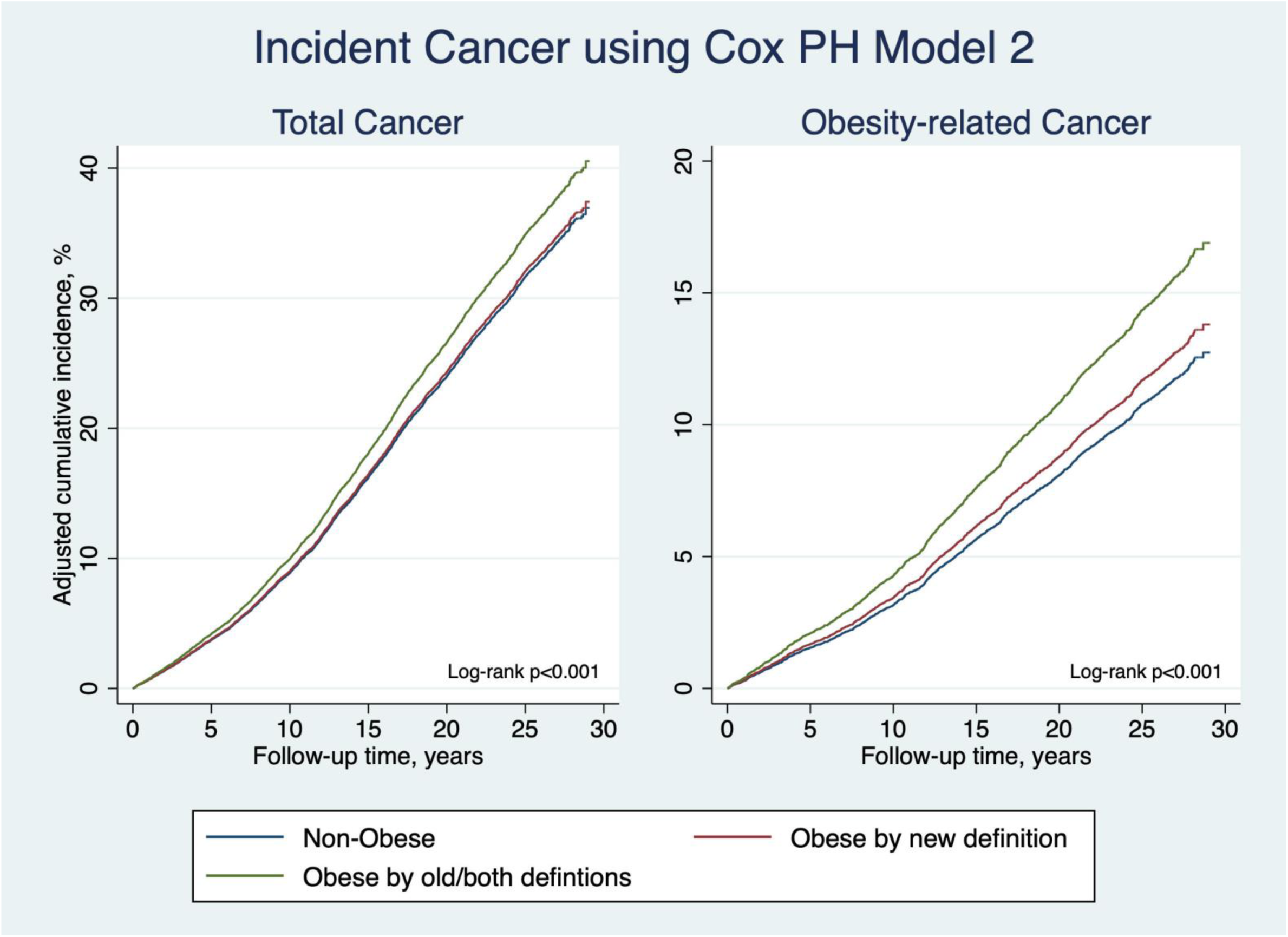
Total and obesity-related cancers events over follow-up period stratified by obesity definition.

**Table 3.**
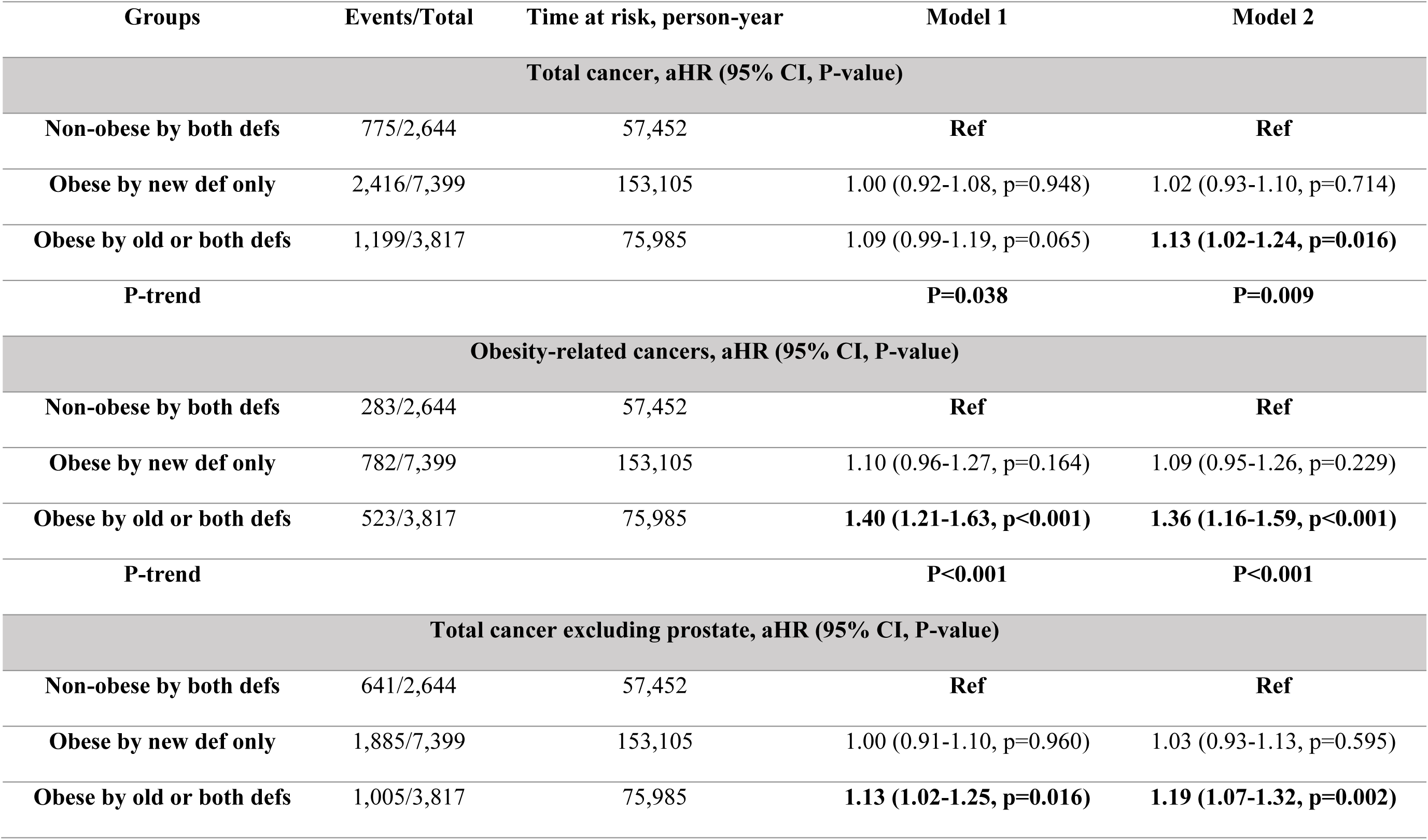
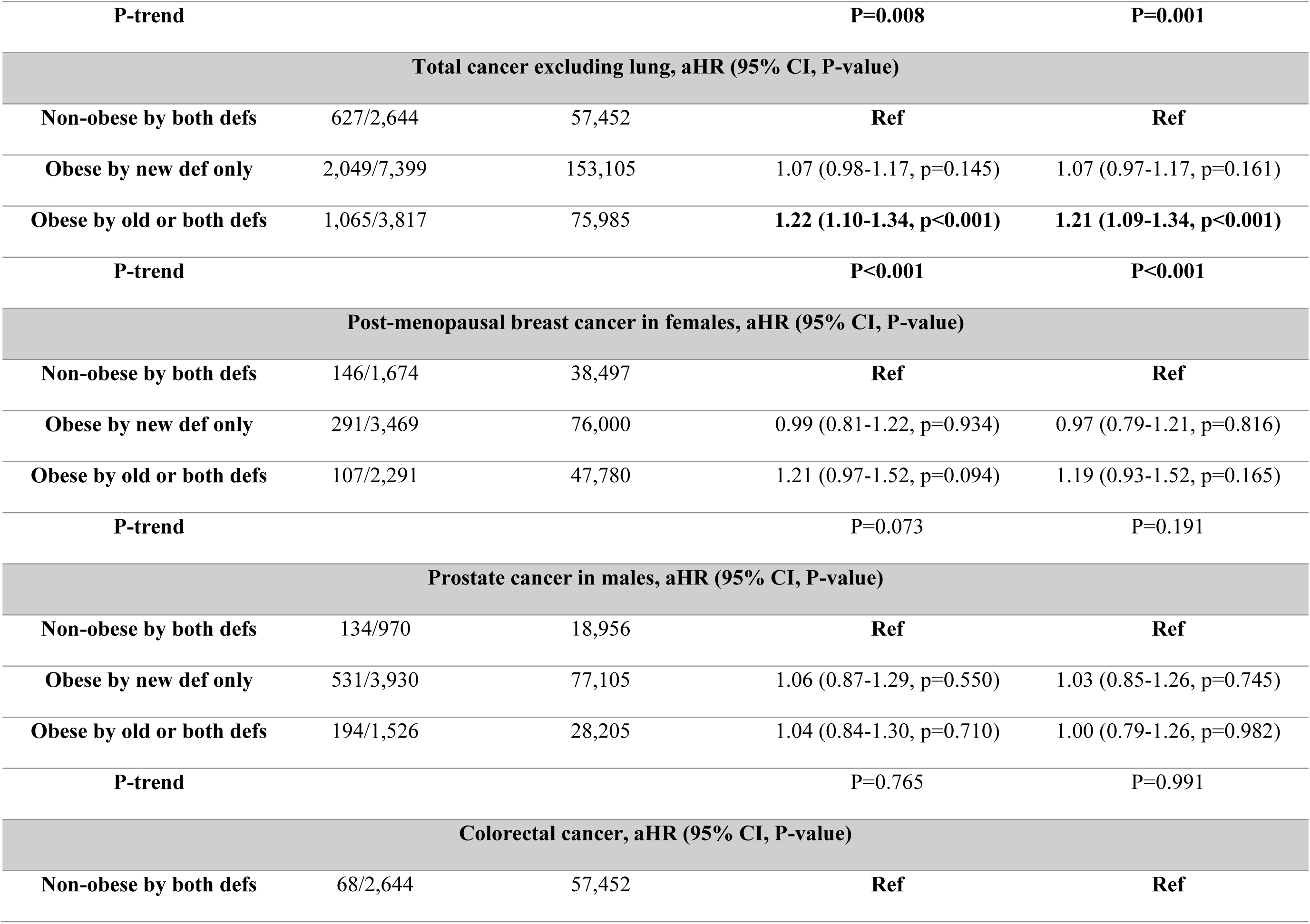
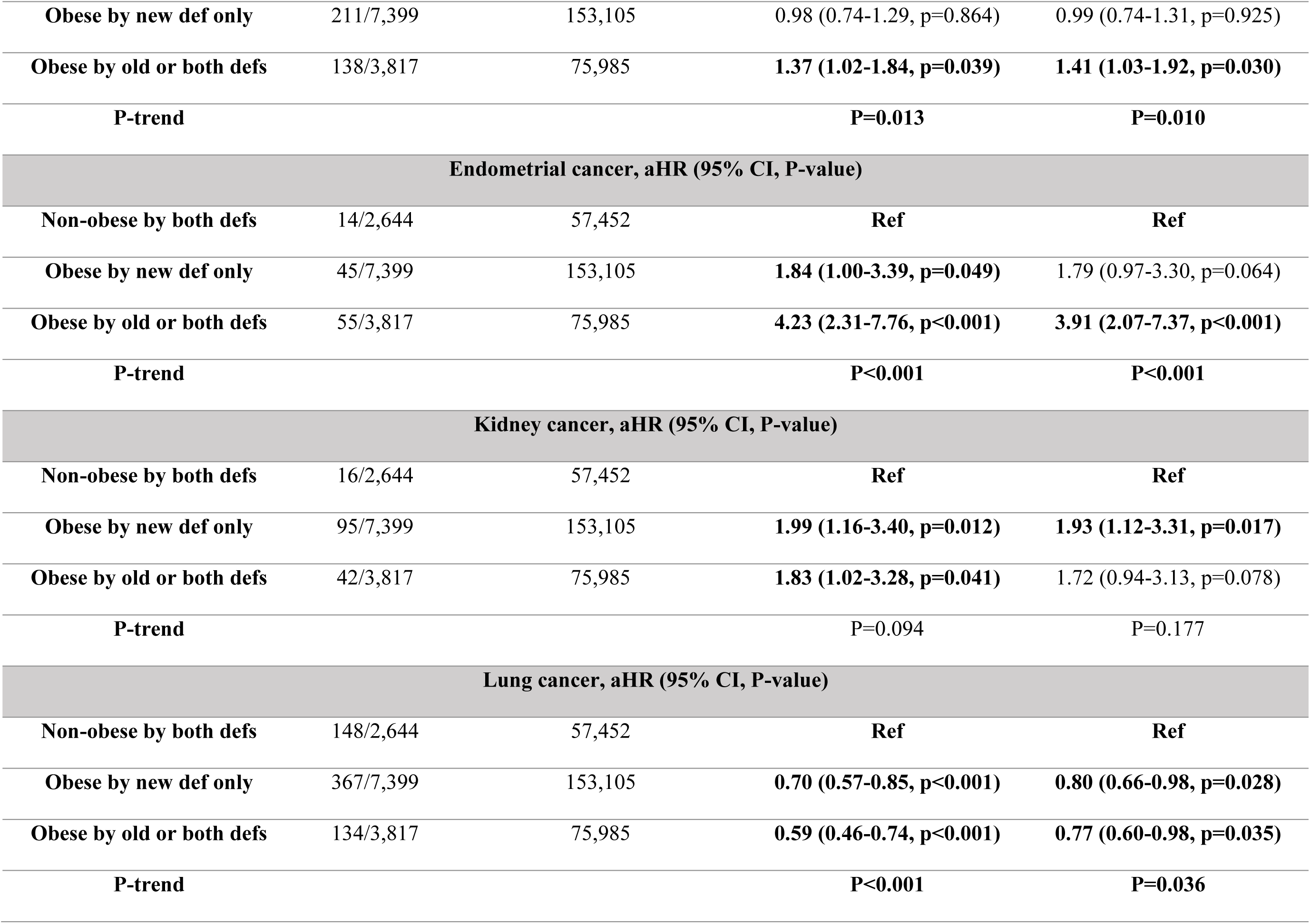
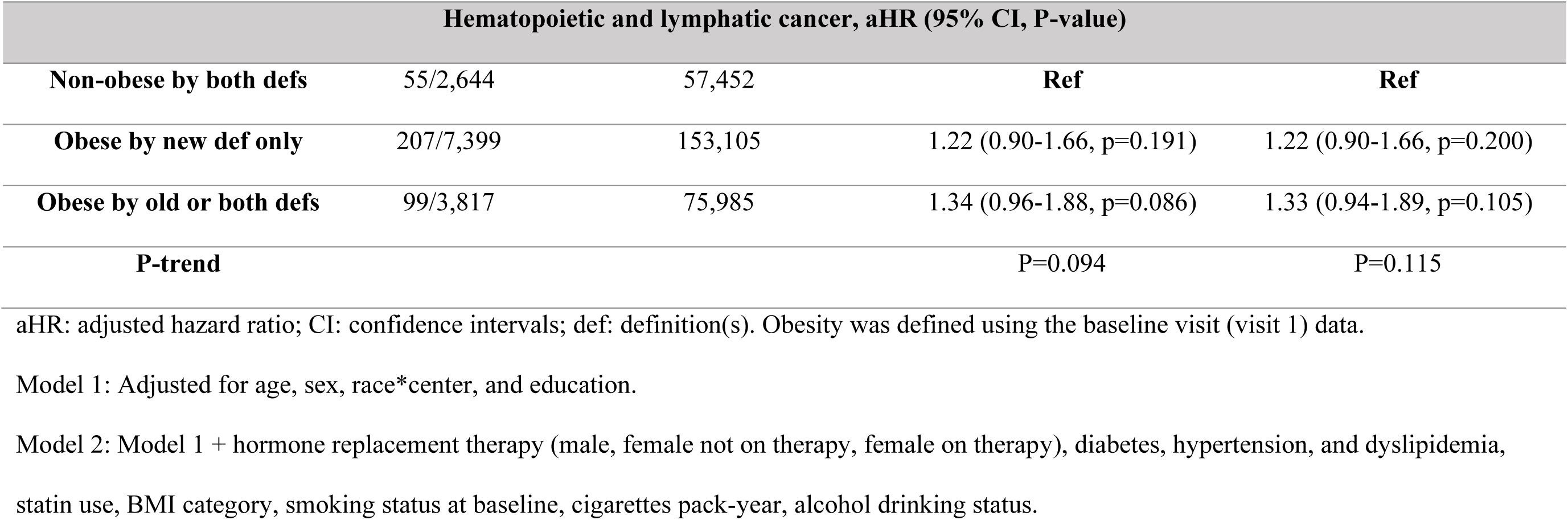
Association between obesity definitions and the incidence of cancer, 13,860 men and women in ARIC, 1987-2015. Bold text indicates significant results.

Using the time-updated obesity, similar patterns of findings were observed for total cancer (Model 2: old/both definitions aHR 1.12, 95% CI 1.01-1.25; new-only definition: aHR 1.03, 95% CI 0.93-1.13) and obesity-related cancers (old/both definitions: aHR 1.41, 95% CI 1.18-1.68; new-only definition: aHR 1.12, 95% CI 0.95-1.33), whereby obesity categorized by old/both definitions was associated with a higher risk while obesity categorized by the new-only definition was not **(Table S9)**. Patterns for colorectal, prostate, and lung were unchanged using time-updated obesity definitions compared with using baseline obesity definitions. For endometrial and kidney cancers, the association for time-updated obesity categorized by the old/both definitions remained (aHR 2.24, 95% CI 1.38-3.63), while the association for the time-updated obesity categorized by the new-only definition was attenuated and not statistically significant. For post-menopausal breast cancer (old/both definitions: aHR 1.30, 95% CI 1.01-1.69), and hematopoietic and lymphatic cancers (aHR 1.49, 95% CI 1.01-2.22), associations for obesity by old/both definitions were stronger and statistically significant when using time-updated definitions, while obesity by the new definition remained non-significant **(Table S9)**.

In the subgroup analysis, a similar pattern was noted in both male and female subgroups, where only the individuals meeting the old obesity definition showed higher risk of obesity-related cancers (male: aHR 1.48, 95% CI 1.05-2.09; female: aHR 1.38, 95% CI 1.16-1.65; **Table S10-S11)**. Also, a similar pattern to the overall population was demonstrated in the female subgroup, where both groups of obesity had more total cancers and endometrial and kidney cancers than the non-obese, but those meeting the older definition criteria had significantly higher risk (P-trend p<0.001; **Table S11)**. In the race subgroups, only White individuals demonstrated significant association between both the older definition (aHR 1.44, 95% CI 1.20-1.73) and the newer definition (aHR 1.19, 95% CI 1.02-1.40) of obesity and obesity-related cancers. Similar results were demonstrated in total cancer and endometrial and kidney cancers **(Table S12)**. On the other hand, Black individuals did not show any significant association between either obesity definitions and obesity-related cancers, although they showed lower risk of total cancer, which became non-significant after excluding lung cancer **(Table S13)**. Lastly, both groups who had events or were censored before or after the age of 70 years did not show any significant associations between either obesity definition and obesity-related cancers. In those censored or who had events before the age of 70 years, those classified as obese the old definition had a lower incidence of total cancer, which became non-significant after excluding lung cancer **(Tables S14-S15)**.

Finally, missingness analysis and covariates imputations in both CVD and cancer analysis did not demonstrate any significant deviation from the original results **(Table S16)**.

## Discussion

In this large prospective analysis of the ARIC cohort, we aimed to compare the long-term associations between obesity and incident CVD and cancer, comparing the traditional BMI-based definition of obesity with a newer framework, as suggested by a recent Lancet Commission report, that incorporates measures of central adiposity. Our findings produced several insights: first, those classified as obese by either the old or new definition demonstrated an increased risk of incident total CVD, yet those classified as obese by the new definition showed a weaker association. Similar findings were demonstrated for CHD, stroke/TIA, and HF. Second, in terms of cancer, the traditional BMI-based definition of obesity was associated with a significantly higher risk of total cancer, obesity-related cancers, and colorectal cancer, whereas the newer definition of obesity, which identified an additional 54% of the cohort as obese (for a total of 81% obese in the cohort), was not generally associated with these outcomes aside from endometrial and kidney cancers. This increase in the prevalence of obesity for the new definition was mainly due to inclusion of individuals in the BMI-defined overweight category who also had a large waist circumference. A similar finding was recently published using the All of Us cohort.^24^ Third, we demonstrated that those categorized as obese by the new definition tended to transition back to a non-obesity state over time, in part due to lowering BMI and waist circumference. Fourth, the well-known “obesity paradox” was observed for lung cancer, where both definitions were associated with a lower risk; the new definition of obesity did not appear to capture the aspect of adiposity (central) that is positively associated with lung cancer risk.^9^ Finally, time-updated obesity definition patterns of association were generally similar to baseline for those classified as obese by old/both definitions and by the new-only definition, with a few exceptions by cancer site.

The motivation behind the present investigation stems from the limitations of using BMI as the sole measure of adiposity-related health,^3,4^ which does not account for fat distribution, particularly visceral adiposity, which is known for its detrimental metabolic impact.^4,5,7,25^ The recently published Lancet Commission report highlighted these limitations and advocated for a more inclusive diagnostic framework that moves beyond BMI and includes other anthropometric measures of central adiposity, such as waist circumference (WC), waist-to-hip ratio (WHR), and waist-to-height ratio (WHTR).^3^ While a recent paper examined the prevalence of obesity using the new definition in the National Health and Nutrition Examination Survey (NHANES) and concluded that both definitions showed similar prevalences, they included only one anthropometric measure, and waist circumference.^26^

### Cardiovascular Disease Risk

The current finding that both definitions of obesity were associated with incident total CVD are consistent with the literature linking excess adiposity, particularly central adiposity, with both cardiovascular morbidity and mortality. In the present investigation, we demonstrated that both definitions showed a significant association with CVD; however, the “newly” diagnosed group showed a slightly attenuated risk despite the aim of the new classification to capture high-risk central adiposity. It was also notable that these “newly” diagnosed individuals had more favorable cardiometabolic risk profiles at baseline, with lower BMI and reduced prevalence of hypertension and diabetes. This healthier baseline might explain the somewhat lower, yet significant, CVD risk compared to the traditional definition, which inherently includes individuals with more severe overall adiposity and often a longer duration of adverse metabolic states. Nevertheless, this new definition still captured a significant increase in CVD risk in individuals who are overweight and were previously deemed healthy, highlighting its clinical utility in unmasking that risk at an earlier stage than the traditional definition so that earlier interventions, in particular diet and lifestyle, could be implemented to reduce risk over the long-term. Therefore, it is essential to recognize the advantages of the new definition in mitigating that risk; however, the risk is not comparable to those defined as obese under the older definition.^25^

Similar effects were noted for single cardiovascular outcomes, including CHD and most prominently in HF. Also, after subgrouping sex, race, and median age at the time of diagnosis/censorship, the newer definition still demonstrated significant, yet attenuated, associations with CVD. This further demonstrate how the robust findings and associations demonstrated by this new definition.

### Cancer risk

The findings from the relationship between the traditional and newer obesity definitions and cancer were more distinct. The traditional BMI-based definition was associated with a higher risk of total cancer and obesity-related cancer, which aligns with previous literature addressing obesity and cancer risk.^9^ However, those categorized as obese based on the new definition did not have an increased risk of total or obesity-related cancer. This is a critical finding because it suggests that although being overweight (BMI 25-<30 kg/m^2^) along with central adiposity might be associated with a higher risk of CVD, the association with cancer risk is weak, in contrast to the association between more severe obesity (BMI ≥30 kg/m^2^) and/or a large waist circumference and cancer risk.

As previously discussed, it is possible, that the severity and duration of exposure, differences in the location of the adiposity, and consequent pathways perturbed (e.g., inflammatory, metabolic, sex steroid hormonal) might explain the differences observed in associations for total cancer, obesity-related cancers, and site specific cancers among those who are categorized as obese by the traditional BMI criteria versus the new definition. Individuals with a BMI ≥ 30 kg/m^2^ usually tend to have a longer duration of obesity, and as our data support that they have higher rates of diabetes and hypertension prevalences.^1^

Some prior studies suggested that abdominal obesity might be linked to breast and colorectal cancer development and progression, although the results were inconsistent.^8,10,11^ The observed inverse association between both definitions of obesity and lung cancer is consistent with previous literature that described an “obesity paradox” in some studies of lung cancer.^27–29^ Also, for prostate cancer and post-menopausal cancer, no significant association was observed with either of the obesity definitions. This relationship, in particular, can be complex due to the hormone-sensitive nature of those cancers, which could be influenced by cancer aggressiveness (i.e. for prostate cancer), menopausal status, and hormone receptor status (i.e. for breast cancer).^2,30,31^

### Intermediate risk group

A particularly striking and clinically important finding of the present study was the size of the group defined as obese by the new-only definition. This group constituted a substantial 54% of the ARIC population at baseline, comprising individuals with a BMI of 25-<30 kg/m^2^, a median BMI of 26 kg/m^2^, and high central adiposity, as indicated by waist circumference (WC), waist-to-hip ratio (WHR), and/or waist-to-height ratio (WHTR). Using the traditional definition of obesity with a BMI of ≥30 kg/m^2^, these individuals would have been overlooked or classified as “metabolically healthy obesity” or “metabolically unhealthy obesity” based on their other metabolic risk factors, including, presence of hypertension, high triglycerides, low HDL, or high blood sugar.^25,32,33^

This “newly diagnosed with obesity” group represents the part of the broad obesity spectrum that lies in between the two extremes of (1) the completely normal adiposity by both older and newer definitions, and (2) more severe forms of adiposity as defined by the traditional definition of BMI ≥30 kg/m^2^. This distinct group presents an attenuated risk profile compared with those with a BMI ≥30 kg/m^2^, and a significantly higher risk for CVD compared to those who are non-obese. Furthermore, this distinct group of individuals did not demonstrate a higher risk for cancer compared with those with a BMI ≥30 kg/m^2^. This divergence might suggest that the central adiposity in this BMI range, while detrimental, may not carry the same magnitude or breadth of risk as the more severe forms of obesity with a BMI ≥30 kg/m^2^.

The transitional nature of this group could further our understanding of its characteristics. Our analysis indicates that a notable portion of those individuals transitions back to the non-obese state during follow-up, while a few of them progress to meet the traditional definition of obesity, and the majority remain in the same category. This suggests that this category is relatively stable, an intermediate state for several individuals, but also a dynamic classification with potential for improvement or progression to other states of adiposity.^34^ Thus, labeling this large group of the general adult population as simply “obese”, as suggested by the newer definition, might lead to an overestimation of their absolute risk for specific outcomes and contribute to diagnostic inflation and the risk of medication overutilization or the premature consideration of intensive interventions. This becomes more concerning in high-prevalence settings, such as cardiology clinics, which already manage populations with a high comorbidity burden.

On the other hand, we envision that this newer definition, incorporating central adiposity, appears most valuable as a screening tool in primary care and family medicine clinics to identify individuals with high central adiposity, thereby warranting closer monitoring, targeted lifestyle counseling, and management of associated risk factors. The newer framework is a sensitive tool by nature that was able to identify everyone with a BMI over 30 kg/m^2^ but also was able to identify individuals who could be at higher risk of CVD who were previously overlooked. However, those individuals do not carry the same risk profile as other individuals with BMI over 30 kg/m^2^.^35^

### Strengths and Experimental Considerations

This study has several strengths, which include: (1) using data from a large, well-characterized prospective cohort with a long-term follow-up for incident events; (2) all anthropometric measures and BMI were performed by trained technicians, minimizing self-report bias often encountered in other datasets; (3) very minimal missingness of data in terms of anthropometric measures and other covariates which allowed for detailed analysis of the association between two distinct definitions of obesity and two separate outcomes, while also stratifying by baseline diseases and various site-specific cancers; (4) the longitudinal nature of the various anthropometric measures taken over time, allowed us to consider the update in obesity status over time and incorporate that in the analysis.

However, limitations and experimental considerations must be acknowledged. First, as an observational study, residual confounding cannot be entirely ruled out despite adjustments for a wide range of covariates. Second, for some site-specific cancers and stratified analyses, the sample size and the number of events were small, thus limiting the statistical power. Third, the practical implementation of the newer definition faces challenges. Unlike the abundant use of height and weight to calculate BMI, these measurements, such as waist circumference and hip circumference, are neither routinely collected nor systematically collected in most outpatient or inpatient settings.^5,36,37^ The broader implementation and adoption of such measures would require changes in clinical workflows, the integration of electronic health records, and potentially additional training for healthcare personnel. Fourth, the ARIC study data did not include dual-energy x-ray absorptiometry (DEXA) scans and thus they were not used in the central adiposity definition. However, if available, we can envision an even higher prevalence of obesity, not a lower one. Fifth, the findings may not be generalizable to the entire population, particularly those from different racial/ethnic groups with different body composition patterns and individuals from other settings different from the ARIC study centers. Sixth, in the sensitivity analyses using time-updated obesity definitions, we employed Lexis expansion to split follow-up time by visit intervals. While this approach allowed obesity status to be updated, the irregular visit spacing—particularly the 13-year interval between visits 4 and 5—means that covariates may not fully capture concurrent exposure status throughout follow-up. However, the consistency between our baseline and time-updated analyses suggests minimal bias. Finally, the definitions of “high” central adiposity thresholds (WC, WHR, WHTR) used were based on standard cutoffs; however, optimal risk-stratified cutoffs may vary across different populations.

### Clinical and public health implications

This study’s findings emphasize that obesity is a heterogeneous condition, and its assessment requires more than BMI alone.^3–5^ The newer framework successfully identifies a unique group of individuals, often in the overweight BMI category, who carry an intermediate risk profile, distinct from both non-obese and traditionally obese individuals. Further characterization, including cardiometabolic and inflammatory profiling, is warranted for a better understanding and risk stratification of this group of individuals.

Clinically, our findings suggest that individuals with a BMI between 25 and <30 kg/m^2^ should not be considered uniformly as low risk, but they should be further evaluated for central adiposity (e.g., waist circumference, etc.), despite its practical limitations, which is crucial to identify those who might benefit from proactive monitoring, lifestyle modifications, and risk factor management.^5,36,37^ Efforts to integrate these measurements into primary care are warranted for enhancing screening and targeted prevention.

## Conclusion

This study demonstrated that while both traditional BMI-based and newer obesity definitions incorporating central adiposity are associated with increased risk of CVD, and heart failure in particular, compared to non-obesity, the newer definition of obesity was associated with a comparatively lower CVD risk than the traditional definition. However, these associations might vary in magnitude across the population subgroups. For cancer, only the traditional definition was consistently associated with an increased risk of cancer, and obesity-related cancers in particular. The newer definition identifies a unique large population segment – often those with BMI 25-<30 kg/m^2^ and central adiposity – associated with intermediate CVD risk profile, distinct from those non-obese individuals and those defined as obese by the traditional definition. While integrating central adiposity measures increases the sensitivity to identify an intermediate-risk group for targeted preventive strategies, careful consideration of terminology is certainly needed to avoid potential risk inflation. The choice of obesity definition markedly impacts the observed risk associations, demanding a careful approach in clinical and public health settings.

## Data Availability

Data from the Atherosclerosis Risk in Communities (ARIC) Study are available through established ARIC data sharing procedures. Due to privacy protections, specific datasets used in this analysis may be available upon request through the ARIC Coordinating Center, subject to their data policies and approval. Information about ARIC data access is available at https://sites.cscc.unc.edu/aric/.

## Acknowledgment

The authors thank the staff and participants of the ARIC study for their important contributions. Cancer data were provided by the Maryland Cancer Registry, Center for Cancer Prevention and Control, Maryland Department of Health, with funding from the State of Maryland and the Maryland Cigarette Restitution Fund. The collection and availability of cancer registry data are also supported by the Cooperative Agreement NU58DP007114, funded by the Centers for Disease Control and Prevention. Its contents are solely the responsibility of the authors and do not necessarily represent the official views of the Centers for Disease Control and Prevention or the Department of Health and Human Services.

